# Children’s outdoor play at early learning and child care centres: examining the impact of environmental play features on children’s play behaviour

**DOI:** 10.1101/2025.01.21.25320884

**Authors:** Rachel Ramsden, Ian Pike, Sally Thorne, Mariana Brussoni

## Abstract

Early learning and child care centres are critical settings to support children’s regular, repeated and quality time spent in outdoor play. Gibson’s theory of affordances highlights the importance of the human-environment relationship, emphasizing how children use environmental information to inform their behaviour. This study aims to understand the association between children’s outdoor play behaviour and common environmental play features in early learning and child care outdoor play spaces, through the behaviour patterns of children. Children’s play behaviour was collected via observational behaviour mapping at eight early learning and child care centres in the Greater Vancouver region between September 2021 and November 2022, as part of the PROmoting Early Childhood Outside study. A multivariate logistic regression model examined the association between outdoor play behaviour and environmental play features, via odds ratio and 95% confidence intervals. The results indicate environmental play features, including gardening areas, playhouses, climbing structures and tricycle paths supported increased opportunities for children’s outdoor play. Gardening areas, playhouses, sandboxes, outdoor stages and fixed water features provided opportunities for exploratory play, while climbing structures and trike paths provided opportunities for physical play. Opportunities for diverse forms of play were less realized in dedicated open play areas, with the availability of loose parts and moveable equipment primarily influencing these spaces. The results of this study have important implications for future early learning and child care outdoor space design. Further research should consider children’s dynamic movement and transition between outdoor affordances, and the influence of loose parts on the use of environmental play features.

## Introduction

Early learning and child care (ELCC) environments are a fundamental component of a child’s microsystem [1]. Outside of the home, ELCC settings are the most influential environment in a child’s life for outdoor play to transpire [2,3]. Research has shown that outdoor play in the early years is associated with many health and developmental benefits, including enhanced cognitive, physical, emotional and social development, spatial awareness, motor-skills, and physical activity [4–9]. Despite these benefits, contemporary trends, such as changing neighbourhood landscapes, increased screen time, and the prevalence of structured activities, combined with families’ longer work hours, have led to a reduction in the time children spend engaged in outdoor play [10–13]. Shortcomings in sufficient outdoor play participation are also visible within the ELCC landscape; recent research demonstrates that children routinely do not receive adequate outdoor play opportunities while attending structured settings, such as ELCC centres and primary schools [14–17].

There is a collective perception that children’s play effortlessly occurs in structured environments outside the home [18,19]. When children attend an ELCC centre, it is anticipated that they naturally play through adult-directed activities or self-directed autonomy. Parents have expressed that child care serves a critical role to provide free play opportunities for young children, in particular for working families where they feel time for play is limited at home [20]. The existing discrepancy between societal expectations of children’s play in ELCC centres and the actual state of children’s outdoor play in these settings can be partially attributed to the assumption that children will play anywhere, without consideration of the environmental opportunities and affordances that are provided. While there is robust evidence supporting the notion that there are attributes of an outdoor play space that can encourage, or hinder, children’s participation in outdoor play [21–26], ELCC centres are routinely planned, designed, and built with minimal consideration of children’s play preferences [27]. This is inarguably true in densely populated urban areas, where the availability of green space typically shapes the design of outdoor ELCC infrastructure [28,29]. While Canada includes a wealth of natural green spaces [30], prescriptive licensing regulations and competing space requirements contribute to reduced outdoor play opportunities in ELCC centres [31], a challenge echoed in other geographic contexts [29,32–34]. Children’s outdoor play space design practices frequently adhere to a ‘spaces left-over after planning’ approach [32], where the placement and design of outdoor play areas are an afterthought to the adjacent indoor infrastructure. The significance of providing high-quality outdoor play spaces for children continues to be overlooked, despite the outdoor environment – such as size and diversity of materials – being consistently identified as a critical barrier to meeting minimum outdoor play time standards for ELCC centres in Canada [35] and globally [29,34,36,37].

### Outdoor Play and ELCC Environments

Outdoor spaces at ELCC centres influence the frequency, duration, quality and type of play [38–40], yet they may not provide the optimal settings necessary to encourage and facilitate outdoor play opportunities [34,41]. Artificial surfacing, plastic and metal materials, and single-purpose equipment currently dominate ELCC centres in urban contexts [42]. They are frequently devoid of vegetation and challenging play equipment. Artificial, prefabricated environments are common due to the perception that natural environments are more expensive, harder to maintain and offer more risk for injury [42,43]. In addition, societal fears on injury and risk contribute to the safety-focused design of ELCC spaces that routinely prioritizes fixed play elements, such as traditional play structures [44].

Previous research has examined how outdoor play space design supports affordances for play [26,39,45–47]. Thoughtful combinations of natural and built environment elements, as well as purposeful circulation between outdoor play features, can enhance children’s outdoor play and support a greater use of the space [38,48]. Research has also demonstrated that naturalized environments with plentiful opportunities to engage with natural plants and materials support children’s development and well-being [49–51]. Uneven and sloping terrain can enhance children’s physical development, including balance and agility [43,52–54]. In addition, bushes and trees can create shelters that support opportunities for imaginative play [55,56]. The inclusion of loose parts in outdoor play spaces can also encourage imaginative play [55,56]. In comparison to traditional playgrounds with prefabricated structures, natural and nature-based play spaces have been shown to provide more opportunities for diverse forms of children’s play to take place [22]. While there is existing evidence on outdoor play environments that support children’s outdoor play, many studies have identified a gap in the research on the effects of specific natural and built design features and how they are used [57–60].

### Diversity of Play as Evidence for Play Value

Preschool-aged children (3-5 years) have the developmental capacity to participate in many forms of play, including functional play [61,62], constructive play [61,62], social play [63] and dramatic play [64], as well as structured games [65]. Refshauge et al. [66] proposed that affordances for play should therefore be climb-able, jump-on-able, run-able, balance-able, sing-on-able, imagine-able, touch-able, move-able, mould-able and construction-able. When considering the diversity of play opportunities available for young children, researchers have focused on how best to maximize participation in a diverse array of play behaviour. That is, when children are afforded more opportunities for diverse forms of play to transpire, the play value of an environment is enhanced [22,67]. While the typology for categorization of children’s play may differ within existing literature [62,68–70], there is a consistent central theme that multiple forms of play support the outdoor play experience.

### Theoretical Framework

Gibson’s theory of affordances focuses on the relationship between an organism (individual) and its environment [71]. It emphasizes how individuals perceive and interact with the world around them based on the opportunities, or affordances, that the environment offers. Kyttä [72] further specifies that affordances are perceived directly by the user without the need for complex cognitive processing. The environment, or a component of the environment, provides the source of information and triggers a behavioural response, such as a form of play behaviour.

Affordances are dynamic and context-dependent; an element of the environment can change for a child depending on the situation, abilities or goals [73,74]. In addition, affordances can hold a different meaning and potential for each child based on the child’s knowledge, experience, strength, size, skills and preferences [73]. In the context of children’s play, a slide may serve as a sliding apparatus on a sunny day, while it may also become a water race track for toy boats on a rainy day. Outdoor affordances are not limited to physical actions but can elicit imagination and a realm of possibilities [72–74].

As outdoor play environments are planned, designed and built by adults, Gibson’s theory of affordances supports the important question, ‘are children using these spaces as intended?’. Through this theoretical lens, this study conceptualizes natural and built environmental features by their opportunities for meaningful interaction by children [73]. In the context of this study, it is particularly important to acknowledge that children will perceive affordances differently than adults. Adults may design an outdoor space with an intended use, however, it is up to the child to make use of outdoor elements as they see fit. Considering a transactional approach to study the human-environment relationship, this paper seeks to assess outdoor ELCC environments and associated affordances, through the behaviour patterns of children.

### Research Purpose

This paper delves into the intricate relationship between outdoor environments and the play experiences of young children. While many studies have reinforced the positive impact of play on child development, health and wellbeing, the influence of outdoor ELCC environments on young children’s play remains an evolving area of research. Using an affordance-based theoretical framework, this study seeks to understand how children perceive their environment, not as a collection of objects, but as a set of opportunities for action as demonstrated through their play behaviour. The objective of this research is to analyze common environmental play features’ associations with children’s outdoor play. Through this research, we seek to answer two research questions:

1. What is the association between outdoor ELCC play space features and children’s outdoor play; and
2. How does children’s actual use of outdoor affordances in their ELCC environment align with the intended use, as designed by adults?

## Materials and Methods

### Umbrella Study

This study is part of the PROmoting Early Childhood Outside (PRO-ECO) randomized controlled trial that aims to increase the amount of time children spend participating in outdoor play in ELCC. The PRO-ECO study evaluates the PRO-ECO intervention, which includes a built environment modification to outdoor play spaces, within eight ELCC centres in the Greater Vancouver region (Canada). Data for this study were collected as part of the PRO-ECO trial from September 2021 to November 2022 at three time points: Fall 2021 (Time 1), Spring 2022 (Time 2), and Fall 2022 (Time 3). A detailed study protocol for the PRO-ECO study was published elsewhere [75].

### Inclusion/ Exclusion Criteria

Eight ELCC centres delivering full-day group child care and participating in the PRO-ECO trial were included in this study. Children in this study were aged 2 to 6 years and attending a participating ELCC centre. Continuous recruitment of children occurred between August 2021 and November 2022 through ECE’s at each ELCC centre who distributed consent forms and initiated face-to-face conversations with families. Children were excluded from the study if written parental consent was not received. Over the course of the study, a total of 217 children participated in the PRO-ECO trial and are included in this study’s dataset.

### Study Sites

The eight participating centres were located in 3 different cities of the Greater Vancouver region. Three of the ELCC centres were located above-grade and had rooftop outdoor play spaces. The remaining five centres had indoor and outdoor spaces located at-grade. All centres had outdoor spaces that were directly adjacent to their indoor space. Most of the participating centres had outdoor spaces between 170-230 m^2^ However, two centres had larger outdoor spaces (Centre B & Centre C). All centres had a climbing structure, with most (6/8) containing a traditional fabricated play structure with a climbing wall, stairs and a slide. Concrete surfacing was present in all eight participating centres and often used as a gross motor tricycle path. Each of the participating centres had gardening areas, - while some (5/8) had at-grade garden beds, most had raised concrete planters (7/8). Further information on the eight participating centres can be found in Table 1 and Fig 1.

**Figure 1:**
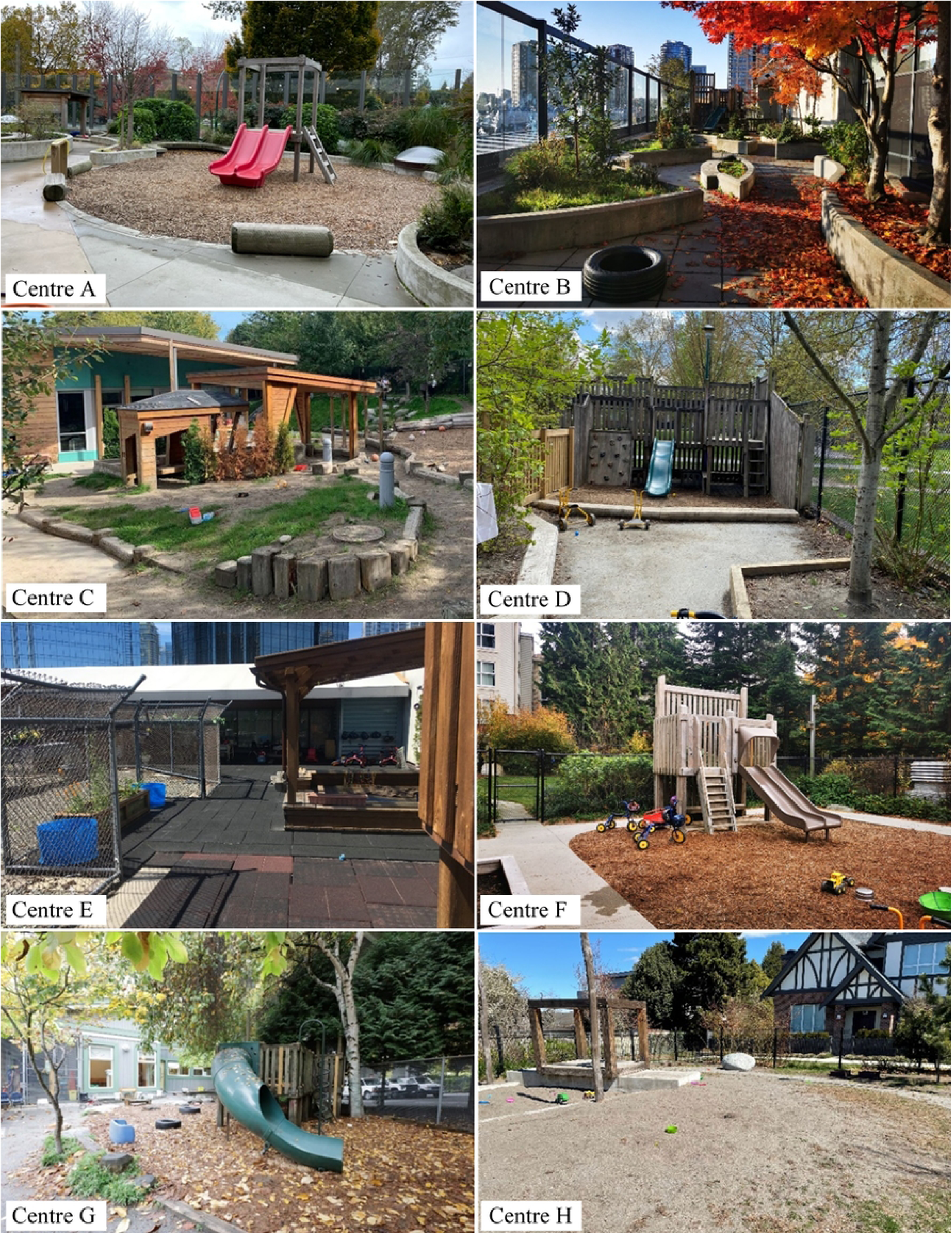
Descriptive photos of participating ELCC centres (n=8) at Time 1

**Table 1:** Characteristics of participating ELCC centres (n=8).

| Attribute | Centre |
| --- | --- |

|  | Centre A | Centre B | Centre C | Centre D | Centre E | Centre F | Centre G | Centre H |
| --- | --- | --- | --- | --- | --- | --- | --- | --- |
| Approximate size (m<sup>2</sup>) | 335 | 171<sup>a</sup> | 754 | 196 | 222 | 270 | 173 | 207 |
| Grade<sup>b</sup> | Above-grade | Above-grade | At-grade | At-grade | Above-grade | At-grade | At-grade | At-grade |
| Climbing structure | Play structure with slide, wood stumps | Play structure with slide | Climbing hill with slide, boulders, intertwined climbing logs* | Play structure with slide, boulders, wood stumps | Play structure with slide | Play structure with slide, balance logs, wood stumps | Play structure with slide, boulders | Wood cubes, balance logs, wood stumps |
| Gross motor path | Concrete tricycle path | Concrete tricycle path | Concrete tricycle path | Concrete tricycle path | None | Concrete tricycle path | Concrete tricycle path | Concrete tricycle path; gravel path |
| Surfacing materials | Concrete, natural soil/ dirt, mulch, artificial turf, wood decking* | Concrete, rubber, wood decking | Concrete, natural soil/ dirt, mulch | Concrete, natural soil/ dirt, mulch, rocks | Concrete, rubber, wood decking | Concrete, mulch, natural soil/ dirt | Concrete, mulch, natural soil/ dirt | Concrete, mulch, natural soil/ dirt, gravel |
| Gardening area | Raised concrete planters | Raised concrete planters | Garden beds | Garden beds, raised concrete planters | Raised concrete planters | Garden beds, raised concrete planters | Garden beds, raised concrete planters | Garden beds, raised concrete planters |
| Water feature | None | None | None | Rain catchers* | None | None | Water pump (moveable)* | Water pump & trough (fixed) |
| Sandbox | Yes | Yes | Yes | Yes | No | Yes | Yes | Yes |
| Mud kitchen | No | Yes | No | No | No | No | No | No |
| Table area | No | Yes | Yes | No | Yes | No | No | Yes |

\* These items were modified/added as part of the PRO-ECO intervention before Time 2 data collection.
<sup>a</sup> This centre increased in size minimally at Time 3 due to an extension to their outdoor space as part of the PRO-ECO intervention.
<sup>b</sup> Grade refers to the ground relationship to the building. Above grade indicates an outdoor play space above ground level. At-grade indicates an outdoor play space at ground level.

### Data Collection

This study used observational behaviour mapping (OBM) to measure children’s outdoor play behaviour in association with their outdoor ELCC space during dedicated outdoor times. This approach strives to understand how an environment supports movement behaviours by mapping, recording, organizing, displaying, and analyzing geographically located data [76,77]. OBM completes recordings of participant movement and behaviours within a given environment to determine how participants use a designated space [77]. In this study, OBM was used to collect approximately 200, 15-second observational behaviour points at each ELCC centre and data collection time point. Data were collected over 30 days at each timepoint: October 2021 - December 2021 (Time 1); April 2022 - June 2022 (Time 2); October 2022 - December 2022 (Time 3). These data collection periods were selected as part of the PRO-ECO study protocol [75] and strived to maintain seasonal similarities between all data collection timepoints.

Within the OBM protocol, data were collected on children’s outdoor play behaviour using the Tool for Observing Play Outdoors (TOPO) developed by Loebach and Cox [68]. Environmental characteristics of the outdoor play space were captured from each centre’s base map and additional variables, including gender and loose parts interaction, were integrated into the data collected within the OBM protocol. The reliability of the OBM method was measured by the degree of interrater reliability and agreement, using weighted κ and intraclass correlation coefficients [78,79]. All OBM points were collected at the centre-level, where children’s play was assessed in relation to their outdoor space. This is in contrast to data collected at the child-level, where play behaviour is assessed in relation to individual children. OBM was implemented through centre-level data collection approaches to facilitate understanding of children’s outdoor play at each participating ELCC centre. All researchers participated in training sessions on the OBM methodology prior to field work. A κ value of 0.918 (agreement=95.9%) was achieved prior to beginning data collection. In addition, a 10% sample of data at each time point was double coded from recorded video observations to further ensure interrater reliability and agreement.

### Outcome Variable

Children’s outdoor play behaviour was coded using the expanded version of the TOPO [68]. The TOPO measures children’s play behaviour through validated categories of 8 play types and 1 non-play type, along with their corresponding subtypes (Table 2). For this study, we did not code any digital play behaviour, and therefore, this TOPO play type category was not present within our dataset. A new dichotomous variable, *play participation,* was derived from the TOPO play types to understand differences in *play* vs *non-play* behaviour. The TOPO categories of physical play, exploratory play, imaginative play, play with rules, bio play and expressive play were categorized as *play*. The TOPO categories of restorative play and non-play were combined to create the *non-play* category because restorative play was often paired with non-play activities, specifically resting and onlooking. We did not see a high number of reading or retreat play. As up to three TOPO codes could be assigned to each play observation, additional rules were determined to categorize play observations that were coded as non-play or restorative play and another play type. Further details of this process have been previously reported [80].

**Table 2:** Tool for Observing Play Outdoors (TOPO) developed by Loebach and Cox [68].

| Play Type | Description |
| --- | --- |
| Physical Play | Includes gross motor, fine motor, vestibular and rough and tumble. |
| Exploratory Play | Includes sensory, active and constructive. |
| Imaginative Play | Includes symbolic, sociodramatic and fantasy. |
| Play with Rules | Includes organic and conventional. |
| Bio Play | Includes plants, wildlife and care. |
| Expressive Play | Includes performance, artistic, language and conversation. |
| Restorative Play | Includes resting, retreat, reading and onlooking. |
| Non-play | Includes self-care, nutrition, distress, aggression, transition and other. |

### Primary Explanatory Variable

The primary explanatory variable, environmental play feature, was derived from each ELCC centre’s base map. Centre base maps were created at each data collection time point to provide an overview of the environment at each participating centre. Base maps included information on topography, ground surface and environmental play features. Environmental play features were categorized for this study by adapting categories of common outdoor affordances used in previous research [81,82], and considering shared features across the eight participating centres. The final environmental play feature variable included: sandbox, tricycle path, gardening area, outdoor stage, fixed equipment-playhouse, fixed equipment-climbing structure, fixed-equipment-water feature, and open area. These categories showcase that participating centres primarily contained environmental play features that were fabricated specifically for play and there were limited natural play elements present, such as large climbing trees or boulders. In the case where wooden stumps or logs were present in the outdoor play space, these features were moveable and captured within the loose parts variable also collected through the OBM protocol. Fig 2 displays an example of Centre A’s base map with assigned environmental (play) feature areas, while Fig 3 showcases Centre A’s base map with observational behaviour points for Time 1 overlayed.

**Fig 2:**
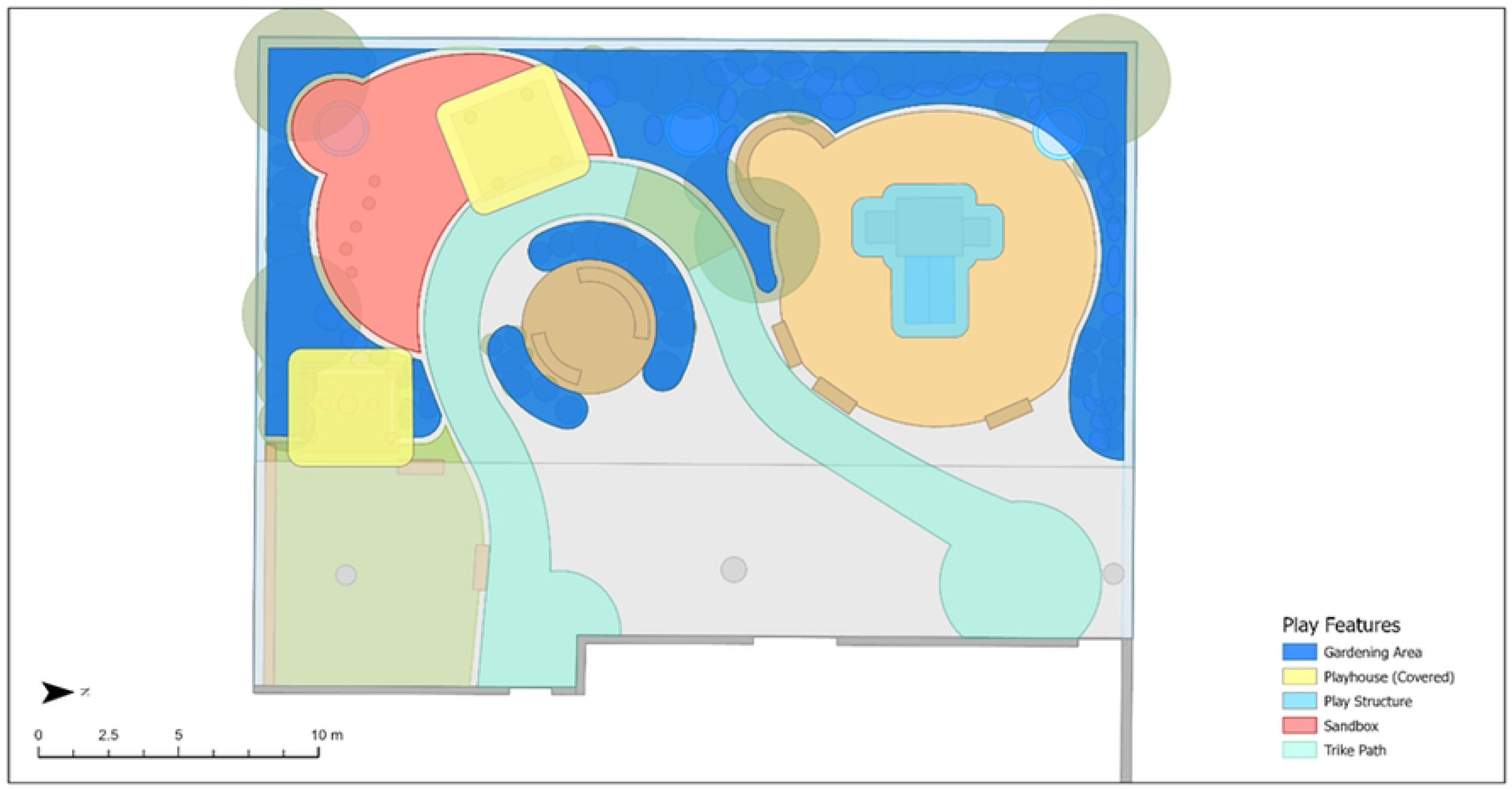
Centre A base map with assigned outdoor play feature areas (non-highlighted areas are open areas).

**Fig 3:**
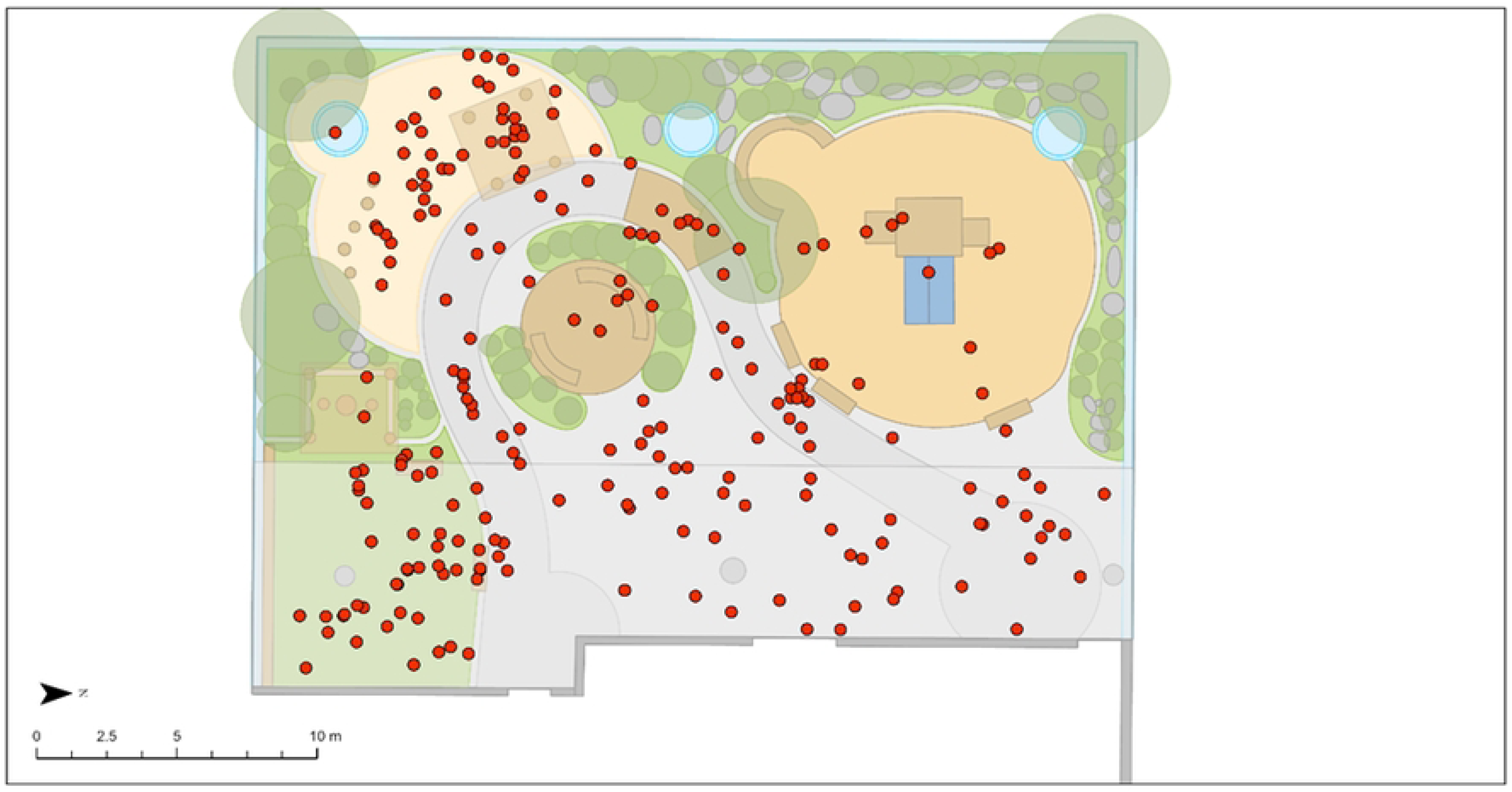
Centre A base map with all observational behaviour points overlayed (Time 1).

### Potential Confounders and Risk Factors

Sociodemographic factors and play factors with known associations with children’s play behaviour were also collected as part of the OBM protocol. Existing research identifies a relationship between children’s outdoor play behaviour and gender [83–85], temperature and weather conditions [83,86–89], ground topography [53,54] and loose parts use [53,56]. Gender was collected during observations and recorded based on how the child presented using potential visible gender markers, as outlined elsewhere by Loebach et al. [90]. Data on temperature and weather conditions were recorded (www.timeanddate.com/weather) and matched to the day and time of data collection. Temperature was included as a continuous variable (°C). Weather conditions were categorized into higher-level categories (cloudy, no rain; raining; sunny).

### Analysis

The analysis for this study sought to understand environmental play features associated with: 1) play participation (vs. non-play participation); and 2) the different play behaviour types captured through the TOPO (Table 2). Univariate analyses were conducted to assess crude associations between play participation and environmental play features, and all covariates. In addition, univariate analyses were completed to compare crude associations between play behaviour type and environmental play features. The primary analysis included a multivariate logistic regression model to obtain an adjusted total effect measuring the association between environmental play feature and the dichotomous outcome variable, *play participation*, via odds ratio (OR) and 95% confidence intervals (CI).

A secondary analysis considered each TOPO play category as a separate play type (physical play, exploratory play, imaginative play, bio play, play with rules and expressive play). The TOPO categories of non-play and restorative play were not included in this analysis as they did not align with this study’s definition of outdoor play. Each of the TOPO play types was looked at independently and the same regression model used for the primary analysis was run for each play type as a binary outcome variable. For both analyses, known confounders and risk factors identified a priori within the literature were included within the model. Multicollinearity was assessed using Variance Inflation Factors (VIF). Statistical analyses were performed using R-4.2.2.

## Results

### Study Observations

A total of 5,213 outdoor observations (play and non-play observations) were collected across the eight participating ELCC centres at three data collection timepoints. Outside of open areas, where observations most frequently occurred (49.1%), tricycle paths (17.6%%), sandboxes (12.0%) and climbing structures (9.4%) were most regularly used by children among all observations (play and non-play). Children were observed most often in low topography terrain (89.9%), while steep (1.2%) and uneven terrains (8.9%) were less commonly observed across observations. This can be attributed to the low prevalence of steep and uneven terrain present across the eight participating ELCC centres. The majority of recorded observations occurred in cloudy weather conditions (62.0%) and included the use of loose parts (75.3%). Boys were more prevalent in this study’s observations (58.6%) than girls (41.4%).

Across all outdoor observations (n=5,213), children were observed participating in play 80.7% of the time (n=4,206). Children’s participation in play was most likely to occur in open areas (46.3%), tricycle paths (17.3%), sandboxes (13.3%) and fixed climbing structures (10.2%). Play observations occurred most frequently on low topography terrain (89.2%), in cloudy weather conditions (62.4%), and with the use of loose parts (79.3%). Non-play observations were most common in open areas (61.0%) and on tricycle paths (18.8%). Table 3 presents the overall frequency distribution for our dataset, stratified by play and non-play participation type.

**Table 3:** Descriptive sample characteristics of all outdoor observations (n=5,213), stratified by play participation type (play or non-play).

| Variable | Stratified by Play Participation Type |  |  | All Observations |
| --- | --- | --- | --- | --- |
|  | Non-Play | Play | p-value |  |
| Number of Play Observations [n (%)] | 1,007 | 4,206 |  | 5,213 |
| Environmental Play Feature [n (%)] |  |  | <0.001 |  |
| Open Area | 614 (61.0) | 1,948 (46.3) |  | 2,562 (49.1) |
| Gardening Area | 26 (2.6) | 219 (5.2) |  | 245 (4.7) |
| Fixed Equipment - Playhouse | 39 (3.9) | 240 (5.7) |  | 279 (5.4) |
| Fixed Equipment - Climbing Structure | 61 (6.1) | 431 (10.2) |  | 492 (9.4) |
| Sandbox | 69 (6.9) | 558 (13.3) |  | 627 (12.0) |
| Outdoor Stage | 7 (0.7) | 49 (1.2) |  | 56 (1.1) |
| Tricycle Path | 189 (18.8) | 727 (17.3) |  | 916 (17.6) |
| Fixed Equipment - Water Feature | 2 (0.2) | 34 (0.8) |  | 36 (0.7) |
| Topography [n (%)] |  |  | <0.001 |  |
| Low | 938 (93.1) | 3,750 (89.2) |  | 4,688 (89.9) |
| Steep | 4 (0.4) | 59 (1.4) |  | 63 (1.2) |
| Uneven | 65 (6.5) | 397 (9.4) |  | 462 (8.9) |
| Weather Conditions [n (%)] |  |  | 0.001 |  |
| Sunny | 202 (20.1) | 944 (22.4) |  | 1,146 (22.0) |
| Cloudy, no rain | 607 (60.3) | 2625 (62.4) |  | 3,232 (62.0) |
| Raining | 198 (19.7) | 637 (15.1) |  | 835 (16.0) |
| Temperature (Mean (SD)) | 8.57 (3.33) | 9.10 (3.13) | <0.001 | 9.00 (3.18) |
| Gender (girl) [n (%)] | 444 (44.1) | 1,715 (40.8) | 0.06 | 2,159 (41.4) |
| Loose Parts Use (yes) [n (%)] | 588 (58.4) | 3,337 (79.3) | <0.001 | 3,925 (75.3) |

The most frequent play types children participated in were physical play (76.5%) and exploratory play (43.9%). Among all physical play observations (n=3,217), 46.1% occurred in open areas, 18.4% occurred on tricycle paths, 12.9% occurred in a sandbox and 11.1% occurred on climbing structures. Playhouses (5.4%), gardening areas (4.2%), water features (0.7%) and stages (1.2%) were less commonly used environmental features when participating in physical play. Observations where exploratory play was coded occurred most often in open areas (41.7%), a sandbox (22.8%) and a tricycle path (14.0%).

While open areas were the most frequently used spaces among all types of play, a large portion of bio play observations occurred in gardening areas (29.6%), including raised planters and at-grade gardening areas. In addition, bio play observations were observed often on uneven terrain (26.0%), which contrasts the low frequency of all outdoor observations (8.9%) occurring on uneven terrain. All play types saw higher frequencies of participation among boys, in comparison to girls. While most play types involved the use of loose parts, play with rules did not have a large proportion of loose parts use (43.8%), in comparison to all outdoor observations (75.3%). Exploratory play (94.6%) and bio play (87.4%) had the highest frequency of loose parts use among all play types.

Weather conditions contributed to different frequencies of play participation, with bio play more likely to take place at higher temperatures (mean: 10.03 Celsius), and less likely to occur when it was raining (9.9%) in comparison to sunny (23.8%) or cloudy (66.4%) weather conditions. Play with rules observations were also less likely to occur in rainy conditions, whereas exploratory play observations were frequently seen in rainy weather (24.6%). Table 4 provides an overview of the frequency of environmental play feature usage and associated covariates, by play behaviour type.

**Table 4:** Descriptive sample characteristics of all outdoor play observations (n=4,206), stratified by play behaviour type.

| Variable | Play Behaviour Type |  |  |  |  |  |
| --- | --- | --- | --- | --- | --- | --- |
|  | Physical Play | Exploratory Play | Imaginative Play | Play with Rules | Bio Play | Expressive Play |
| Number of Play Observations | 3,217 | 1,848 | 404 | 185 | 223 | 891 |
| Environmental Play Feature [n(%)] |  |  |  |  |  |  |
| Open Area | 1,483 (46.1) | 771 (41.7) | 178 (44.1) | 96 (51.9) | 94 (42.2) | 495 (55.6) |
| Gardening Area | 134 (4.2) | 92 (5.0) | 20 (5.0) | 8 (4.3) | 66 (29.6) | 37 (4.2) |
| Fixed Equipment - Playhouse | 175 (5.4) | 136 (7.4) | 30 (7.4) | 1 (0.5) | 0 (0.0) | 55 (6.2) |
| Fixed Equipment - Climbing Structure | 356 (11.1) | 109 (5.9) | 62 (15.3) | 20 (10.8) | 5 (2.2) | 83 (9.3) |
| Sandbox | 415 (12.9) | 422 (22.8) | 50 (12.4) | 6 (3.2) | 9 (4.0) | 86 (9.7) |
| Outdoor Stage | 39 (1.2) | 33 (1.8) | 7 (1.7) | 0 (0.0) | 0 (0.0) | 9 (1.0) |
| Tricycle Path | 593<br>(18.4) | 258 (14.0) | 56 (13.9) | 54<br>(29.2) | 48<br>(21.5) | 123<br>(13.8) |
| Fixed Equipment -<br>Water Feature | 22 (0.7) | 27 (1.5) | 1 (0.2) | 0 (0.0) | 1 (0.4) | 3 (0.3) |
| Topography [n(%)] |  |  |  |  |  |  |
| Low | 2,884<br>(89.6) | 1,650<br>(89.3) | 366 (90.6) | 171<br>(92.4) | 157<br>(70.4) | 816<br>(91.6) |
| Steep | 48 (1.5) | 12 (0.6) | 6 (1.5) | 4 (2.2) | 8 (3.6) | 9 (1.0) |
| Uneven | 285<br>(8.9) | 186 (10.1) | 32 (7.9) | 10<br>(5.4) | 58<br>(26.0) | 66 (7.4) |
| Weather Conditions<br>[n(%)] |  |  |  |  |  |  |
| Sunny | 730<br>(22.7) | 397 (21.5) | 90 (22.3) | 48<br>(25.9) | 53<br>(23.8) | 191<br>(21.4) |
| Cloudy, no rain | 2,016<br>(62.7) | 1,106<br>(59.8) | 257 (63.6) | 122<br>(65.9) | 148<br>(66.4) | 573<br>(64.3) |
| Raining | 471<br>(14.6) | 345 (18.7) | 57 (14.1) | 15<br>(8.1) | 22<br>(9.9) | 127<br>(14.3) |
| Temperature [mean(SD)] | 9.03<br>(3.16) | 9.32<br>(3.12) | 9.14<br>(3.06) | 8.83<br>(3.23) | 10.03<br>(2.74) | 8.91<br>(3.11) |
| Gender (girl) [n(%)] | 1,277<br>(39.7) | 715 (38.7) | 179 (44.3) | 72<br>(38.9) | 99<br>(44.4) | 398<br>(44.7) |
| Loose Parts Use (yes)<br>[n(%)] | 2,555<br>(79.4) | 1,749<br>(94.6) | 311 (77.0) | 81<br>(43.8) | 195<br>(87.4) | 637<br>(71.5) |

<sup>a</sup> Play observations can be coded as (up to) 3 play type categories and therefore, the sum of observations across play types exceeds the sample size (n=4,206)

### Association between Play Participation and Environmental Play Features

The results of the multivariate logistic regression model examining the association between play participation and environmental play features is presented in Table 5. In comparison to open areas, all investigated environmental play features, except for outdoor stages, were significantly associated with children’s play participation. When adjusting for topography, weather conditions, temperature, gender and loose parts interaction, fixed water features (OR:6.30, 95% CI=1.48, 26.76), play structures (OR:3.01, 95% CI= 2.24, 4.04) and gardening areas (OR:2.68, 95% CI=1.75, 4.10) had the highest odds of play participation. Steep slopes (OR:4.17, 95% CI=1.48, 11.75), uneven terrain (OR:1.61, 95% CI=1.21, 2.15), temperature (OR:1.04, 95% CI=1.02, 1.06) and loose parts interaction (OR:3.01, 95% CI=2.58, 3.51) were significantly associated with children’s play participation within the adjusted model. In addition, raining weather conditions were significantly associated with decreased likelihood of play participation (OR: 0.73, 95% CI=0.58, 0.92). These findings indicate that environmental play features beyond outdoor open areas provide significant play worth to children’s outdoor play. Further analysis of the type of play afforded by each environmental play feature is warranted.

**Table 5:** Univariate and multivariate logistic regression results (OR, 95% CI) examining the association between play participation and environmental play feature.

| Variable | Play Participation |  |
| --- | --- | --- |
|  | Unadj. OR<sup>a</sup><br>[95% CI] | Adj. OR<br>[95% CI]<sup>b</sup> |
| Environmental Play Feature |  |  |
| Open Area | Ref | Ref |
| Gardening Area | 2.65 [1.75;4.03]*** | 2.68 [1.75;4.10]*** |
| Fixed Equipment - Playhouse | 1.94 [1.37;2.75]*** | 1.86 [1.30;2.67]*** |
| Fixed Equipment - Climbing Structure | 2.23 [1.68;2.96]*** | 3.01 [2.24;4.04]*** |
| Sandbox | 2.55 [1.95;3.33]*** | 2.19 [1.67;2.87]*** |
| Outdoor Stage | 2.21 [0.99;4.90] | 1.55 [0.69;3.48] |
| Tricycle Path | 1.21 [1.01;1.46]* | 1.23 [1.02;1.49]* |
| Fixed Equipment - Water Feature | 5.36 [1.28;22.37]* | 6.30 [1.48;26.76]* |
| Topography |  |  |
| Low/ No Slope | Ref | Ref |
| Steep Slope | 3.69 [1.34;10.18]* | 4.17 [1.48;11.75]** |
| Uneven Terrain | 1.53 [1.16;2.00]** | 1.61 [1.21;2.15]** |
| Weather Conditions |  |  |
| Sunny | Ref | Ref |
| Cloudy, no rain | 0.93 [0.78;1.10] | 0.90 [0.75;1.08] |
| Raining | 0.69 [0.55;0.86]*** | 0.73 [0.58;0.92]** |
| Temperature | 1.05 [1.03;1.07]* | 1.04 [1.02;1.06]*** |
| Gender |  |  |
| Boy | Ref | Ref |
| Girl | 0.87 [0.76;1.00] | 0.90 [0.78;1.04] |
| Loose Part Interaction |  |  |
| No | Ref | Ref |
| Yes | 2.74 [2.36;3.17]*** | 3.01 [2.58;3.51]*** |

Adj. = adjusted; Unadj. = unadjusted; OR = odds ratio; 95% CI = 95% confidence interval
Note: significance level, \*: p<0.05, \*\*: p<0.01, \*\*\*: p<0.001
<sup>a</sup> Unadjusted OR's calculated via 6 univariate logistic regression models examining play participation and each covariate independently.
<sup>b</sup> OR's from multivariate logistic regression, adjusting for topography, weather conditions, temperature, gender and loose part interaction.

### Associations between Play Behaviour Type and Environmental Play Features

Table 6 outlines the results of the adjusted multivariate logistic regression model examining each play behaviour type and environmental play features. Unadjusted results for each play type outcome are available in Appendix A. Fixed climbing structures (OR:2.23, 95% CI=1.79, 2.78), sandboxes (OR:1.30, 95% CI=1.08, 1.56) and tricycle paths (OR:1.34, 95% CI=1.14, 1.56) were positively associated with children’s physical play at ELCC in comparison to open areas. Rainy weather conditions (OR:0.76, 95% CI=0.63, 0.92), steep slope terrain (OR:2.01, 95% CI=1.10, 3.65) and the use of loose parts (OR:1.92, 95% CI=1.68, 2.19) were all significantly associated with children’s participation in physical play. Girls were less likely to participate in physical play then boys (OR:0.84, 95% CI=0.75, 0.94).

**Table 6:**
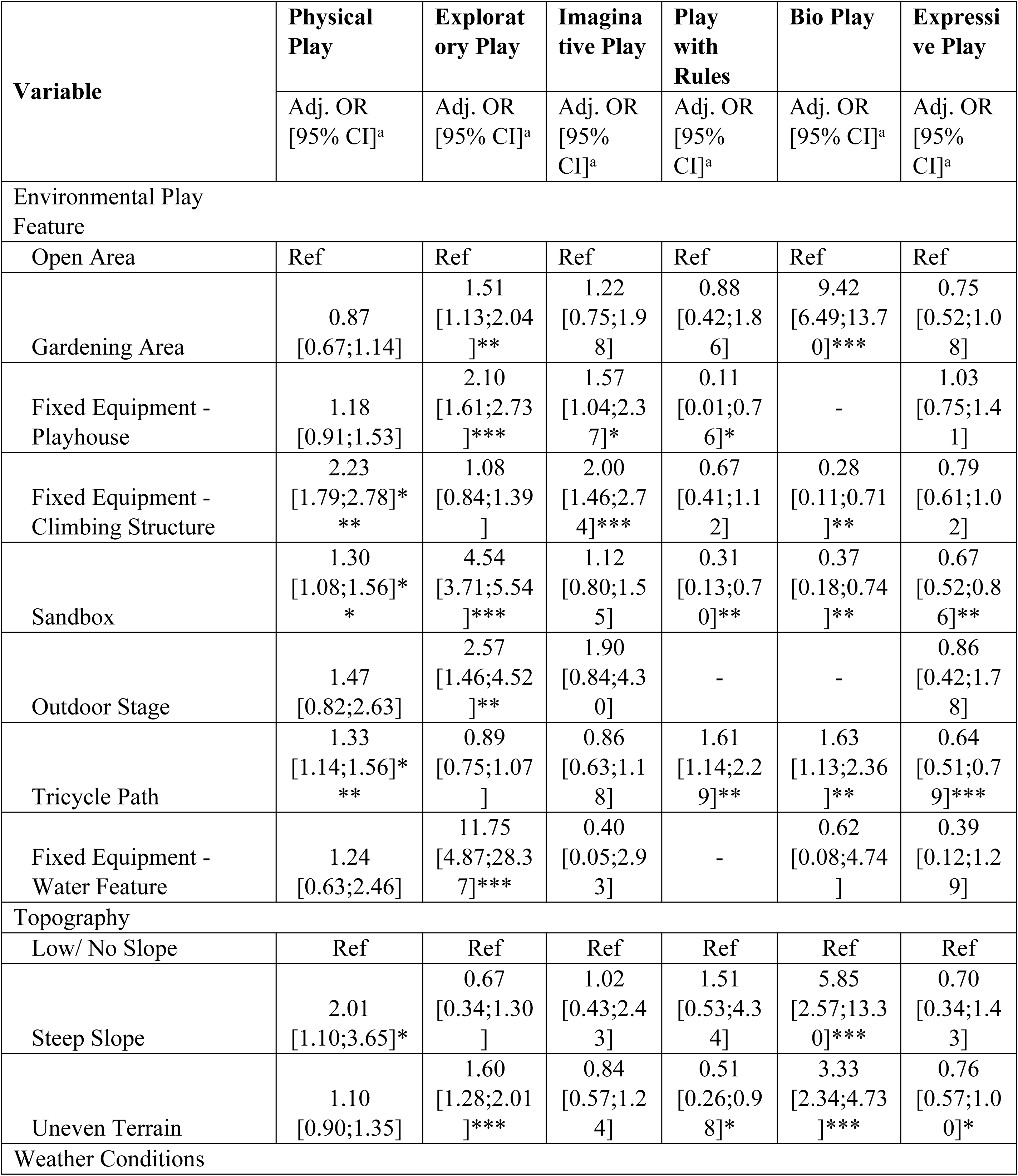
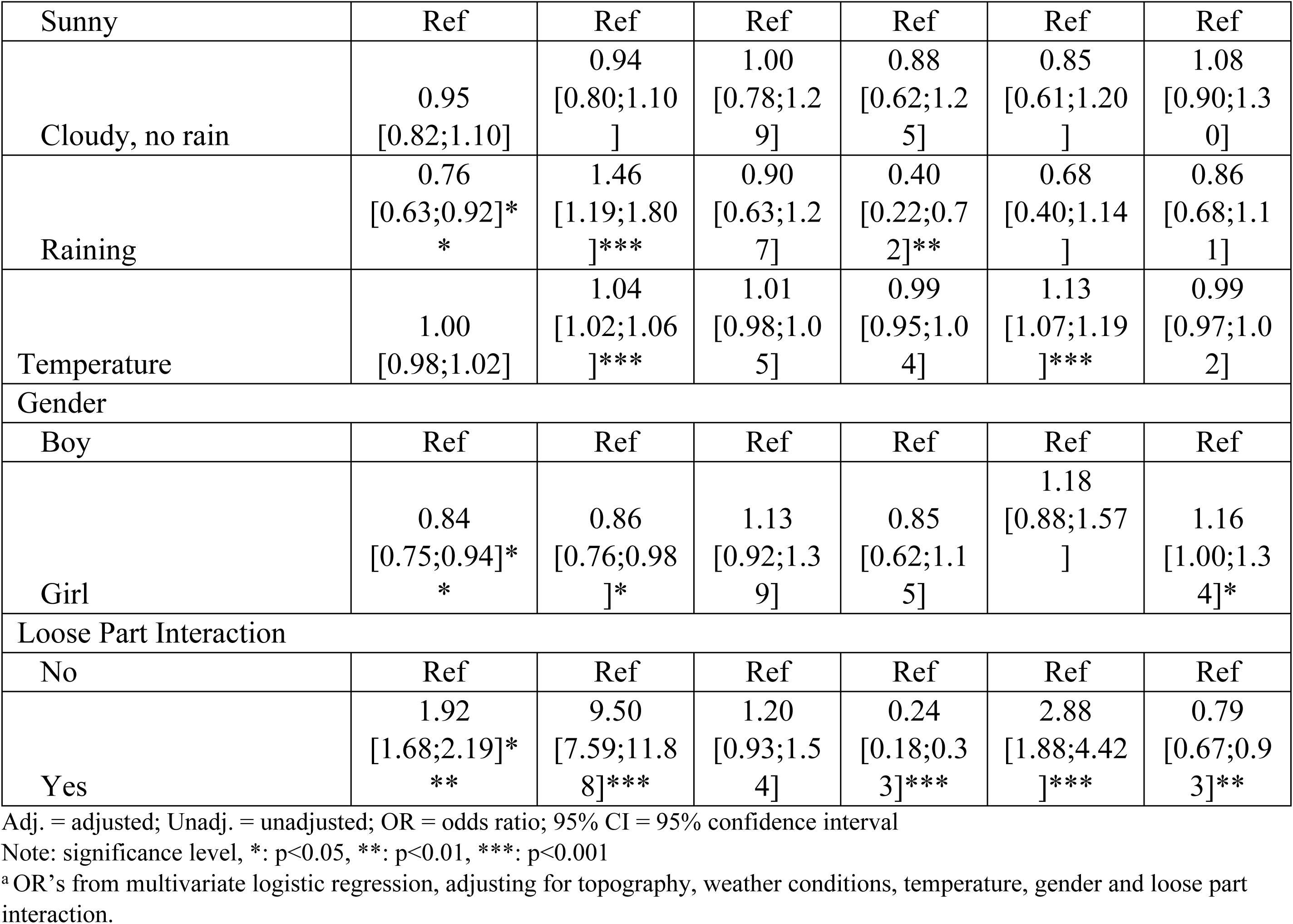
Multivariate logistic regression results (OR, 95% CI) examining the association between play behaviour types and environmental play feature.

Gardening areas (OR:1.51, 95% CI=1.13, 2.04), fixed playhouses (OR: 2.10, 95% CI=1.61, 2.73), sandboxes (OR:4.54, 95% CI=3.71, 5.54), outdoor stages (OR:2.57, 95% CI=1.46, 4.52) and fixed water features (OR:11.75, 95% CI=4.87, 28.37) were significantly associated with children’s exploratory play at ELCC in comparison to open areas. Uneven terrain (OR:1.60, 95% CI=1.28, 201), rainy weather conditions (OR:1.46, 95% CI=1.19, 1.80), temperature (OR:1.04, 95% CI=1.02, 1.06) and loose part use (OR:9.50, 95% CI=7.59, 11.88) were also significantly associated with exploratory play participation within the adjusted model. Girls were less likely to participate in exploratory play then boys (OR:0.86, 95% CI=0.76, 0.98).

Fixed playhouses (OR: 1.57, 95% CI=1.04, 2.37) and climbing structures (OR:2.00, 95% CI=1.46, 2.74) were positively associated with children’s imaginative play at ELCC in comparison to open areas. Gender, weather conditions, temperature and loose parts interaction were not significantly associated with children’s participation in imaginative play. Children’s participation in imaginative play was negatively associated with uneven terrain (OR:0.51, 95% CI=0.26,0.98), rainy weather conditions (OR:0.40, 95% CI=0.22, 0.72) and the use of loose parts (0.24, 95% CI=0.18, 0.33). Tricycle paths (OR:1.61, 95% CI=1.14, 2.29) were significantly associated with increasing children’s play with rules in comparison to open areas. In contrast, playhouses (OR:0.11, 95% CI=0.01, 0.76) and sandboxes (OR:0.31, 95% CI=0.13, 0.70) were negatively associated with children’s participation in play with rules activities. Due to low case numbers, associations between play with rules and outdoor stages and fixed water features was not possible to calculate. Gender was not significantly associated with participation in play with rules.

Gardening areas (OR:9.42, 95% CI=6.49, 13.70) and tricycle paths (OR:1.63, 95% CI=1.13,2.36) were significantly positively associated with children’s bio play in comparison to open areas. Climbing structures (OR:0.28, 95% CI=0.11, 0.71) and sandboxes (OR:0.37, 95% CI=0.18, 0.74) were negatively associated with bio play opportunities. In comparison to low slope terrain, steep slopes (OR:5.85, 95% CI=2.57, 13.30) and uneven terrains (OR:3.33, 95% CI=2.34, 4.73) were significantly associated with increased bio play participation. In addition, interactions with loose parts increased the odds of children’s participation in bio play (OR:2.88, 95% CI=1.88, 4.42). Diverse weather conditions were not associated with bio play; however, bio play did increase in higher temperatures (OR:1.13, 95% CI=1.07, 1.19). There were no significant gender differences in bio play participation.

In comparison to open areas, sandboxes (OR:0.67, 95% CI=0.52, 0.86) and tricycle paths (OR:0.64, 95% CI=0.51, 0.79) were negatively associated with children’s participation in expressive play. There were no significant positive associations with expressive play and environmental play features, indicating that open areas may provide benefits for expressive play participation. Uneven terrain (OR:0.76, 95% CI=0.57, 1.00) and the use of loose parts (OR:0.79, 95% CI=0.67, 0.93) were negatively associated with participation in expressive play. Girls were significantly more likely to participate in expressive play than boys (OR:0.79, 95% CI=0.67, 0.93).

## Discussion

This study aimed to understand children’s play behaviour associations with common environmental play features located across eight urban ELCC centres in the Greater Vancouver region. Findings from this analysis reveal that diverse outdoor play features, including playhouses, climbing structures, sandboxes, tricycle paths and fixed water features, offer significantly more play participation for children than open areas. In addition, gardening areas, playhouses, climbing structures and tricycle paths were environmental play features that offered a positive association with more than one play behaviour type, indicating that these features provided affordances for play diversity.

At each of the participating ELCC centres, open areas represented the largest surface area and were the most frequented areas by children within their outdoor play space. However, open areas did not provide a diversity of outdoor play opportunities in comparison to other examined environmental play features. Similarly, Refshauge et al. [45] found that children’s participation in non-play activities frequently took place in open gathering and open space settings. Open areas within this study may have offered minimal affordances for play based on the way in which the space was set up for children to engage with. As these areas do not have manufactured equipment or designated purposes, it is up to the educators and children themselves to visualize and use the space as they see fit. The available provocations set up in the spaces will guide the behaviours of children [91]. In addition, these spaces are fluid and may change due to weather conditions, educator and staffing resources and the children in attendance on a given day.

This study demonstrated increased expressive play opportunities in open areas in comparison to sandbox and tricycle path areas, however, other play behaviour types were not significantly enhanced in open areas. Opportunities for children’s expressive play behaviour may be enhanced in open areas due to the design of these spaces that are often conducive to social conversations and artistic avenues, such as table areas. This may also be where children feel the most comfortable dancing or performing due to the availability of space to move. While open spaces often provide the largest surface areas for movement, this study demonstrated that tricycle paths were more likely to be used to facilitate physical play in comparison to open areas. Open spaces provided expressive play opportunities, including dancing and performing, but tricycle paths were more frequently used for gross motor activities, such as running, skipping and riding wheeled toys.

The results of this study align with previous research examining outdoor affordances for play among young children [26,45–47]. Sandboxes, climbing structures and tricycle paths were observed to be among the most frequented outdoor affordances within the observations recorded in this study. Similarly, Refshauge et al. [45] found that the most utilized outdoor affordances for play at public playgrounds were fixed play structures, sand play settings, open and gathering spaces, water play settings, playhouses and play ships, and play equipment including spinners, swings and slides. While the results of the Reshauge et al. study are specific to public park settings and include older children, they provide similar insights into the outdoor features that children gravitate towards for their play choices. Further, Holmes and Procaccino [47] observed preschool-aged children outdoor play preferences within ELCC centres. Their results highlighted riding toy areas and sandbox areas as frequently used outdoor play spaces [47].

Tricycle paths were positively associated with the most play behaviour types in this study, including physical play, play with rules and bio play. In these areas, children often chased one another, creating rules for new games and gathering loose parts found in nature nearby. Tricycle paths often provided the only opportunities for children to play at speed, including running or using wheeled equipment. This aligns with existing research demonstrating a strong association between tricycle paths and children’s increased physical activity levels when at ELCC centres [92–94]. Many of the rules-based games involved running or moving fast, and the tricycle path was often identified by educators and children as the preferred area where it was safe to participate in these activities. In addition, many of the organic games that children created involved mobile equipment such as tricycles and scooters, and the tricycle path offered a smooth surface where these games could occur. However, tricycle paths were limited to very specific activities and provided stagnant affordances, where children commonly participated in walking, running or pedaling wheeled equipment. These spaces were negatively associated with expressive play even though tricycle path spaces were among the largest environmental play features in the outdoor play spaces.

Climbing structures were common outdoor play features present across the participating ELCC centres in this study, with all eight centres having a climbing apparatus of some form. These structures were positively associated with children’s physical play opportunities, and provided affordances for climbing, balancing and other gross motor activities. Wishart et al. [46] similarly found that balancing activities, a component of physical play, most frequently occurred on fabricated structures. Previous evidence [26,45] outlines that functional play, such as running, jumping, spinning, and climbing, is commonly observed on play structures, spinning equipment, swings and slides. In addition, recent research outlines children’s physical activity levels as positively influenced by fixed play equipment and playground structures [93–96]. Although accelerometry is often used to measure physical activity levels, Cosco et al. [81] found associations between children’s physical activity and outdoor ELCC features, including open areas, sand play settings, pathways and fixed play equipment, using behaviour mapping.

Exploratory play encompasses play involving sensory play, constructive play or active manipulation of objects. This form of play can also be described as constructive or manipulative play in the literature [54,97]. Among the examined environmental play features, gardening areas, playhouses, sandboxes, outdoor stages and fixed water features, provided opportunities for exploratory play. Many of these features contain sensory components, including soil, sand or water that allow children to manipulate or visually explore their environmental surrounding, a common characteristic of exploratory play. Refshauge et al. [45] found that constructive play was frequently observed in sand and water play settings. Wishart et al. [46] described that within the traditionally designed playscape, manipulative play routinely occurred in sandpits or open yard areas and most often involved the use of loose parts.

While sandboxes were among the most utilized environmental play features in our study, they provided limited affordances for diverse play types. The sandbox is often designed as a ‘catch-all’ spot where all children can locate an activity they enjoy in this space. It also is designed to facilitate groups of children to engage in individual or group play. However, our study showed that sandbox elements are only positively associated with exploratory play. Children were less likely to participate in bio play, expressive play and play with rules in sandboxes than in comparison to open areas. This indicates that the play opportunities offered by a sandbox may be limited and do not resonate with all children, or children may use this play feature in an inconsistent way. In addition, exploratory play is often related to solitary or pair activities, rather than participation in larger groups. Play with rules and expressive play are often group play behaviour activities and entice more cooperative play. Children did not use the sandbox to facilitate games or social play, and therefore, these spaces may not provide consistent and repeated opportunities for diverse play behaviours.

Playhouses were positively associated with exploratory and imaginative play, providing opportunities for children to create sociodramatic activities involving loose parts, such as pretending to bake muffins with mud in a fantasized kitchen. Playhouses were often situated adjacent to sandboxes and provided additional opportunities for explorative play, such as sand and water activities. These fixed structures were often used to facilitate creative interactions with natural elements and peers, including creating a drive-through grocery store, and mixing water and sand to create new recipes. Children often used playhouses for the purposes of using sand and water in an undercover location. Refshauge et al. [45] also demonstrated that dramatic play, similar to imaginative play, was most often observed on play ships or playhouses. This aligns with our findings where imaginative play was positively associated with playhouses and climbing structures. Wishart et al. [46] observed that children’s dramatic play often involved the use of multiple outdoor spaces and affordances, such as pretending to be horses galloping across an imaginary landscape. Recent research by Cetken-Aktas and Sevimil-Celik [26] found that dramatic play was most likely to occur when fixed equipment encompassed a theme or imaginative component, such as a ship, a truck or a house, and in enclosed play spaces, such as under climbing structures or within playhouses.

In the present study, fixed water features had minimal play type associations, with only exploratory play significantly increased in these areas, indicating that there were less opportunities for diverse and robust play in these spaces among our sampled observations.

Children within our study appeared to be intentionally playing with natural materials only near or on designated gardening areas, resulting in many outdoor play features providing minimal affordances for bio play participation. In contrast, gardening areas provided opportunities for exploratory and bio play, supporting children to plant, explore nature or utilize soil and garden materials to construct. Gardening areas are often designed to facilitate exploratory play, where children have sensory opportunities to touch and feel natural materials [98]. This study highlights that gardening areas were the only environmental play feature that offered significant opportunities to participate in both exploratory and bio play, which are important to children’s interaction with nature.

A summary of the adjusted associations between play behaviour type and environmental play feature showcase important findings to guide future design of outdoor ELCC spaces (Table 7).

**Table 7:** Summary of outdoor play type and environmental play feature associations.

| Play Behaviour Type | Environmental Play Feature Associations (in comparison to open areas) |  |  |  |  |  |  |
| --- | --- | --- | --- | --- | --- | --- | --- |
|  | Gardening Area | Playhouse | Climbing Structure | Sandbox | Outdoor Stage | Tricycle Path | Water Feature |
| Physical |  |  | + | + |  | + |  |
| Exploratory | + | + |  | + | + |  | + |
| Imaginative |  | + | + |  |  |  |  |
| Play with Rules |  | - |  | - |  | + |  |
| Bio Play | + |  | - | - |  | + |  |
| Expressive Play |  |  |  | - |  | - |  |

### Intended vs. Actual Use of Outdoor Affordances

Each of the outdoor play spaces at participating ELCC centres was created by adults, including landscape architects, educators and urban planners. It is important to consider the discrepancies between the play design intention and the actual use of these outdoor spaces by children, observed within this study. These results provide important considerations for outdoor ELCC play space design recommendations.

The results from our study indicate that open areas were not significantly associated with most play behaviour types; however, open spaces are often described by educators as preferred spaces for large body movements to facilitate rough and tumble or rambunctious play. Open areas are routinely designed to provide circulation spaces for physical play (moving from space to space, accessing and entering different spaces, running, skipping, walking, etc.). Even with large amounts of space available in open areas, children were observed participating in physical play opportunities more frequently at climbing structures and on tricycle paths. While further research is required to understand the limitations of open outdoor areas at ELCC, the results of this study suggest that open spaces may not entice children to participate in physical play opportunities without the proper provocations in place.

Within this study, the environmental play feature category ‘outdoor stage’ was created to emphasize the significance of these spaces that are often designed in ELCC centres to host performative activities or opportunities, such as singing, dancing, acting or expressive circle times. However, we found that expressive play was not associated with these raised stage areas. Outdoor stages were not commonly found across the eight participating centres and appeared as raised wooden steps at the centres that had this feature. Where they were available, children often used these spaces to jump, balance or participate in other physical play opportunities.

These spaces were not clearly outlined as alluring performative and expressive play opportunities and required the educators to direct or initiate these play forms for them to take place. Future outdoor play space design should consider how raised stages will be used by the educators and if it is necessary to implement a dedicated space, or if this form of play can occur naturally within other play areas or by provocations in open areas.

Fixed water features for children’s use, including water pumps, hoses and water troughs, were often installed to support gardening activities and showcase environmental solutions to sustainability. However, water features were not significantly associated with children’s bio play in our study, indicating that children were not engaging in planting or gardening activities with the use of these water features. Through our observations, we noted that all water features were hard to use without the support of an educator. Children often struggled to navigate the water pumps on their own and required assistance an educator, or at times a peer, to initiate water play. It was also noted that water features were often closed for parts of the year due to weather conditions, such as freezing temperatures. To facilitate bio play opportunities through the use of fixed water features, these pieces of equipment must be accessible to children and easy to navigate without the support of an educator.

Playhouses are normally designed to solicit imaginative play. Built as ‘mini’ versions of a ‘grown-up’ home, they sometimes contain mud kitchens, water tables or features that allow for children to create and conduct their own reality. Children often used playhouses for the purposes of using sand and water in an undercover location. The results of this study showcase that playhouses were frequently used as intended in design, and are important affordances for imaginative and exploratory play to transpire. Playhouses, as well as raised climbing structures, are also created to provide hiding nooks and shelter for children. Surprisingly, our findings did not showcase these structures as significantly associated with play with rules opportunities, such as playing ‘hide and seek’ or similar hiding or tag games. This may be due to the spaces being too open to act as hiding spaces from peers. The playhouses in our study usually had four open sides, consisting of a base and a roof, while the climbing structures often had a minimum of two open sides where educators could view children easily, even if underneath the climber. Enclosed spaces are important components of a child’s outdoor play space to facilitate hiding related to games with peers, offer prospect refuge, or to simply get a reprieve from the concentration of people within the space. It is imperative to consider the enclosed nature of potential hiding and retreat spaces, and how these can be enhanced within play structures to support a diversity of play opportunities.

### Strengths and Limitations

This study is among the first to provide evidence-based information on outdoor environments and associated affordances that support outdoor play opportunities among children aged 2 to 6 in ELCC centres. While previous studies have explored children’s preferences and observed behaviour with natural materials or the macro-design of outdoor spaces [99], this study considers play affordances within a child’s direct microenvironment. This study also considers variation in outdoor play spaces, both geographically and structurally, by capturing data at 8 ELCC centres in different urban communities, and with different sizes and characteristics. In addition, this study collected data at three time points and across various weather conditions to create a robust sample of children’s outdoor play behaviour.

A limitation of this study was the occurrence of the COVID-19 pandemic that impacted the normal operations of participating ELCC centres, including outdoor time and behaviour. Data collection began in mid-2021, when many COVID-19 restrictions were in place in the province, which may have impacted the outdoor behaviour of children or their response to researchers being present in their ELCC spaces. In addition, ELCC centres experienced high staff-turnover during this time and outdoor play practices may have been impacted. To account for the possible limitations due to COVID-19 restrictions, the research team connected with participating ELCC centres frequently and made field notes to address potential changes in normal routines.

## Conclusion

The results of this study provide important findings on the play value of outdoor affordances that are commonly found in urban ELCC centres in the Greater Vancouver region. Open areas are often the largest and most frequented spaces within outdoor play settings at ELCC; however, open areas provided reduced affordances for play in comparison to other measured outdoor play features. Consideration of how outdoor open areas are set up and supported by educators is important to enhance these spaces. These results highlight the importance of considering children’s actual use versus the intended design of play spaces.

Future research should consider the impact of movement across built environment features, considering how children may diversify their play behaviour by using multiple spaces and features of the environment. Furthermore, further research would benefit from considering the role of loose parts as a mediator in the play behaviour and environmental play feature relationship. The results identified within this study provide guidance for outdoor space design within ELCC settings.

## Data Availability

All dataset files will be available from the UBC Research Data Collection database following acceptance.

## Acknowledgements

The authors are grateful to the YMCA GV for their partnership on this study and the members of the Outside Play Lab that supported data collection throughout the PRO-ECO study to inform this manuscript.

